# Clinical signs, symptoms, perceptions and impact on quality of life of patients suffering peri-implant diseases: a university-representative cross-sectional study

**DOI:** 10.1101/2020.06.08.20125229

**Authors:** Mario Romandini, Cristina Lima, Ignacio Pedrinaci, Ana Araoz, Maria Costanza Soldini, Mariano Sanz

## Abstract

**Aim:** To study the symptoms and perception reported by patients with peri-implant diseases, as well as their signs and their potential impact on the oral health quality of life.

**Materials and Methods:** 240 randomly selected patients were invited to participate. As part of the history assessment, the patient OHIP-14Sp was evaluated together with, for each implant, the patient perception regarding the peri-implant health status and the history of pain, spontaneous discomfort, bleeding, suppuration, swelling and discomfort during brushing. As part of the clinical examination, the following potential signs of peri-implant diseases were collected: probing pocket depth (PPD), mucosal dehiscence (MD), extent of BoP, presence of SoP, and visual signs of redness and swelling. Those parameters were analyzed in relation to the actual peri-implant health diagnosis.

**Results:** 99 patients with a total of 458 dental implants were studied. Even in case of peri-implantitis, 88.89% of the implants were perceived by the patients as healthy. The total OHIP-14Sp sum score did not differ in relation to the peri-implant health diagnosis. Increased reports of spontaneous discomfort, bleeding, swelling and discomfort during brushing were observed in presence of disease. However, only a minor proportion of implants with peri-implant diseases presented symptoms. PPD≥6 mm was more frequent in diseased than in healthy implants(p<0.01), while PPD≥8 in pre-periimplantitis/peri-implantitis than in healthy/mucositis implants(p<0.01). Implants with peri-implantitis showed higher MD than implants without peri-implantitis(p<0.01).

**Conclusions:** Peri-implant diseases are in most cases asymptomatic and not perceived by the patients. Despite being unable to accurately discriminate between peri-implant mucositis and peri-implantitis, PPD and MD resulted as the only two clinical signs associated with pre-periimplantitis/peri-implantitis.

## Introduction

Peri-implant diseases are plaque-associated inflammatory lesions occurring in the tissues around dental implants (Berglundh, et al., 2018a; Zitzmann & Berglundh, 2008). They are highly prevalent in patients with dental implants (Derks & Tomasi, 2015; Derks, Schaller, Hakansson, et al., 2016a; Rakic et al., 2018; Rodrigo et al., 2018; Romandini, Cordaro, Donno, & Cordaro, 2019; Vignoletti, Di Domenico, Di Martino, Montero, & De Sanctis, 2019; Wada et al., 2019) and include two main entities: peri-implant mucositis and peri-implantitis. While inflammation of the peri-implant mucosa is a common aspect, the differential feature between the two diseases is the loss of supporting bone, which characterize peri-implantitis (Berglundh, et al., 2018a; Renvert, Persson, Pirih, & Camargo, 2018).

Despite different non-surgical and surgical treatment strategies proposed for treating peri-implantitis, disease resolution is seldom the long-term outcome and even when achieved, recurrence and implant loss may occur (Berglundh, Wennström, & Lindhe, 2018b; Carcuac et al., 2017; Cha, Lee, & Kim, 2019; de Tapia et al., 2019; Figuero, Graziani, Sanz, Herrera, & Sanz, 2014; Heitz-Mayfield et al., 2018; Nart et al., 2020; Ravidà, Saleh, et al., 2020b; Roccuzzo, Layton, Roccuzzo, & Heitz-Mayfield, 2018). In light of this limited predictability, its prevention and early diagnosis and treatment become the most appropriate strategy.

Regular maintenance therapy of patients with dental implants has shown to reduce the risk of peri-implant diseases (Monje et al., 2016; Monje, Wang, & Nart, 2017; Schwarz, Derks, Monje, & Wang, 2018), acting both as a preventive measure and as an occasion for intercepting and treating the diseases in the early stages (i.e. peri-implant mucositis), when their control is easier. However, the actual patient compliance to maintenance recalls is generally low (Amerio et al., 2020; Romandini et al., 2020). In this context, the presence of symptoms could potentially alert patients with peri-implant diseases to seek dental care before the disease is too advanced. However, although the symptoms and patient’s perception to periodontitis have been extensively studied and include gum bleeding during toothbrushing and tooth mobility (Chatzopoulos, Cisneros, Sanchez, Lunos, & Wolff, 2018; Dietrich et al., 2007; Eke et al., 2013), the symptoms and patient perception to peri-implant diseases, as well their impact on oral health related quality of life, have been not thoroughly studied.

Beside the evaluation of patient’ reported symptoms and perceptions, another important aspect of diagnosis is the evaluation of the clinical signs specifically associated to peri-implant diseases. Indeed, despite that an essential component of peri-implantitis diagnosis is the presence of radiographic bone loss, clinical signs should be used to discriminate which patients really need a radiographic evaluation of their implants, thus reducing unjustified radiographic exposures. However, there are few studies that have analysed all the clinical signs associated with peri-implant diseases (Monje, Caballé-Serrano, et al., 2018a; Monje, Insua, et al., 2018b; Ramanauskaite, Becker, & Schwarz, 2018).

It was, therefore, the main objective of this observational study to evaluate the patient’s reported symptoms and perception of their peri-implant health and disease, as well as their potential impact on the oral health related quality of life. Secondarily, this study aimed to study the clinical signs associated with peri-implant diseases.

## Materials and Methods

The present cross-sectional study is reported according to the STrengthening the Reporting of OBservational studies in Epidemiology (STROBE) guidelines (Elm et al., 2007; Vandenbroucke et al., 2007). The research protocol was ethically approved by the CEIC Hospital Clínico San Carlos, Madrid, Spain (19/182-E).

### Sampling procedures

The detailed sampling procedures are reported elsewhere (Romandini et al., 2020). In brief, in order to generalize the results to all the patients who received implants in the Master of Periodontology of the Complutense University of Madrid, we used a complex protocol though a stratified multistage sampling (Sanz & Chapple, 2012; Tomasi & Derks, 2012). Consequently, basing on a sample size calculation, three patients were randomly selected, by computer-generated randomization lists, for each periodontist (or post-graduate student in periodontology) that placed implants in at least 10 patients, from September 2000 to July 2017, in the referred clinic. During each academic year, periodontists who placed implants in less than 10 patients were grouped into a single category and 3 patients were also selected for this group.

The selected patients were invited to participate in the study by telephone calls on the numbers reported in their clinical charts, and if no response, the patient was not discarded until at least five attempts on different days have been made. Only implants with at least one year of loading were evaluated.

### Data collection

The participants who accepted to participate underwent a through data collection process, consisting of four phases: collection of demographic and medical/dental history data, a clinical examination, a radiographic examination and an analysis of their past dental records (detailed description reported in Romandini et al., 2020).

Briefly, the history collection was structured in two steps. The first one (self-reported questionnaire) was based on the completion of written questionnaires by the study participants, after a brief explanation by one interviewer. The self-reported questionnaire also included the oral health impact profile 14 assessment using its validated Spanish version (OHIP-14Sp; León, Bravo-Cavicchioli, Correa-Beltrán, & Giacaman, 2014). The OHIP-14Sp consists of 14 questions including seven domains (functional limitation, physical pain, psychological discomfort, physical disability, psychological disability, social disability, handicap). A Likert-type scale ranging from 0 to 4 (0 = never; 1 = hardly ever; 2 = occasionally; 3 = fairly often; 4 = very often) is used to answer each question. Responses can be summed up in each domain and overall. The total OHIP-14Sp sum score can ranges from 0 to 56, and higher scores indicate a poorer OHRQoL (Graetz, Schwalbach, Seidel, Geiken, & Schwendicke, 2020).

The second step of the history collection (structured interview) was based on a series of standardized questions personally asked by a trained interviewer. Among them, the patients had to answer for each implant, with the aid of a facial mirror to precisely localize it, their perception regarding the peri-implant health status (“Do you think that your implant is in health?”) and the history of pain, spontaneous discomfort, bleeding, suppuration, swelling and discomfort during brushing.

The entire history collection was carried out before any diagnosis of peri-implant status was made. Consequently, when answering the patients were blinded in most cases (i.e. excluding past disease diagnosis) about their peri-implant health status.

The clinical oral examination was carried out by 2 calibrated examiners and included the assessment of patient-, restoration- and implant-related variables. The implant-related examination included the following measurements: probing pocket depth (PPD), mucosal dehiscence depth (Sanz-Martín et al., 2020), bleeding and suppuration on probing (BoP/SoP, within 30s) and presence of visual signs of redness and swelling (not at all, mild, moderate, severe). The peri-implant probing measurements were carried out using a manual UNC-15 periodontal probe (PCP15; Hu-Friedy, Chicago, IL, USA) at 6 sites/implant. The inter-examiner agreement on 10 patients (23 implants) was calculated and resulted in substantial agreement for the deepest PPD (ICC=0.69; p<0.001) and for the presence of mucosal dehiscence (agreement=86.36%; kappa=0.70; p<0.001), moderate agreement for swelling (agreement: 7.27%; kappa=0.57; p<0.01), and only slight agreement for redness (agreement=59.09%; kappa=0.18; p>0.05) (Landis & Koch, 1977). Since the clinical oral examination was carried out before any radiograph was taken, the examiners were unaware in most cases (i.e. excluding past diagnosis) on the peri-implant health status.

Peri-apical radiographs of the included implants were also obtained. The level of marginal bone (BL) was assessed by a single calibrated investigator using a software program (Autocad 2016 TM, AutoDesk Inc., San Rafael, CA, USA) (Flores-Guillen, Álvarez-Novoa, Barbieri, Martin, & Sanz, 2018). One month after the initial evaluation, 50 randomly selected radiographs were re-measured by the same investigator to calculate the intra-examiner agreement (ICC=0.98; 95% CI 0.96-0.99; p<0.001).

### Peri-implant health and diseases case definitions

The following case definitions were used (Romandini et al., 2020; Sanz & Chapple, 2012):

- Peri-implant health: absence of BoP/SoP;
- Peri-implant mucositis: presence of BoP/SoP together with radiographic BL<1 mm;
- Pre-periimplantitis: presence of BoP/SoP and 1 mm≦BL<2 mm;
- Peri-implantitis: presence of BoP/SoP together with radiographic BL ≧ 2 mm.

### Data analysis

All statistical analyses were performed using STATA version 13.1 software (StataCorp LP, College Station, TX, USA). Descriptive characteristics of the population and implants were initially summarized. Patient perception, symptoms and signs were described at implant-level, and their relationship with the peri-implant health status was analysed through multilevel logistic regressions to account for the patient variance (due to the presence of more than 1 implant in most participants), and the relative p-values reported. The OHIP-14Sp was also described at patient-level according to each one of its domains and as sum score, and differences according to the patient peri-implant status were analysed through Kruskal-Wallis test, with the relative p-values reported.

## Results

The sampling strategy resulted in the selection of 240 subjects and 110 of them accepted to participate receiving the examination. From this initial sample, one patient was excluded as only presenting one implant loaded from less than 1 year, while another patient was excluded due to the loss of all the implants. Due to the absence of readable radiographs of all the implants, 9 further patients were excluded from the present analysis, resulting in a total analysed sample of 99 patients. Those 99 patients had a total of 475 implants; however, 2 implants were excluded since were loaded from less than 1 year and 15 implants excluded due to the absence of readable radiographs. Consequently, the present analysis included a total of 99 patients with 458 dental implants with at least 1 year of loading time.

### Descriptive statistics of the study population and implants

Table 1 and table 2 provide descriptive statistics of the study population and implants. Most of the included patients were women (60.61%), currently non-smokers (81.81%), with moderate/severe periodontitis (62.89%) and with a mean age at examination of 63.71 years. Most of the implants were placed in the maxilla (55.24%) and rehabilitated through bridges (58.30%) and screw-retained prosthesis (49.34%). The prevalence of peri-implantitis was 56.57% at patient-level and of 27.95% at implant-level.

**Table 1.** General characteristics of the study population.

|  | Overall<br>(N=99) | Peri-implant<br>health/mucositis<br>(N=12) | Pre-periimplantitis<br>(N=31) | Peri-implantitis<br>(N=56) |
| --- | --- | --- | --- | --- |
| **Age** (years), mean (SD) | 63.71 (9.26) | 58.16 (12.04) | 64.64 (8.11) | 64.39 (8.95) |
| **Gender**, N (%) |  |  |  |  |
| Male | 39 (39.39) | 4 (33.33) | 9 (29.03) | 26 (46.43) |
| Female | 60 (60.61) | 8 (66.67) | 22 (70.97) | 30 (53.57) |
| **Educational Level**, N (%) |  |  |  |  |
| Primary school | 32 (32.32) | 2 (16.67) | 25 (80.65) | 18 (32.14) |
| High school | 26 (26.26) | 2 (16.67) | 2 (6.45) | 14 (25.00) |
| Middle grade | 20 (20.21) | 3 (25.00) | 2 (6.45) | 12 (21.43) |
| University/College | 21 (21.21) | 5 (41.67) | 2 (6.45) | 12 (21.43) |
| **Smoking Status**, N (%) |  |  |  |  |
| Non-smokers | 41 (41.41) | 4 (33.33) | 15 (48.39) | 22 (39.29) |
| Former smokers | 40 (40.40) | 2 (16.67) | 3 (9.68) | 21 (37.50) |
| Current smokers | 18 (18.18) | 6 (50.00) | 13 (41.94) | 13 (23.21) |
| **Marital Status**, N (%) |  |  |  |  |
| Married | 73 (73.74) | 9 (75.00) | 25 (80.65) | 39 (69.64) |
| Widow | 6 (6.06) | 1 (8.33) | 2 (6.45) | 3 (5.36) |
| Divorced | 9 (9.09) | 1 (8.33) | 2 (6.45) | 6 (10.71) |
| Never married | 8 (8.08) | 0 (0.00) | 2 (6.45) | 6 (10.71) |
| Living with unmarried partner | 3 (3.03) | 1 (8.33) | 0 (0.00) | 2 (3.57) |
| **Diabetes Status**, N (%) |  |  |  |  |
| No diabetes | 83 (83.84) | 11 (91.67) | 26 (83.87) | 47 (83.93) |
| Diabetes | 16 (16.16) | 1 (8.13) | 5 (16.13) | 9 (16.07) |
| **Periodontal Status (AAP)**, N (%) |  |  |  |  |
| No/Mild Periodontitis | 27 (27.84) | 5 (41.67) | 13 (43.33) | 9 (16.36) |
| Moderate/Severe Periodontitis | 61 (62.89) | 6 (50.00) | 17 (56.64) | 38 (69.09) |
| Edentulous | 9 (9.28) | 1 (8.33) | 0 (0.00) | 8 (14.55) |
| **Regular Maintenance**, N (%) |  |  |  |  |
| No | 46 (46.94) | 6 (53.33) | 16 (53.33) | 24 (42.86) |
| Less than one/year | 8 (8.16) | 3 (10.00) | 3 (10.00) | 5 (8.93) |
| One/year | 21 (21.43) | 7 (23.33) | 7 (23.33) | 10 (17.86) |
| Two/year | 19 (19.39) | 3 (10.00) | 3 (10.00) | 15 (26.79) |
| Three or more/year | 4 (4.08) | 1 (3.33) | 1 (3.33) | 2 (3.57) |

Total number varies according to missing data for each variable. SD, standard deviation; N, number.

**Table 2.** General characteristics of the study implants.

|  | Overall<br>(N=458) | Peri-implant<br>health (N=39) | Peri-implant<br>mucositis<br>(N=146) | Pre-<br>periimplanti<br>tis<br>(N=145) | Peri-implantitis<br>(N=128) |
| --- | --- | --- | --- | --- | --- |
| **Jaw, N (%)** |  |  |  |  |  |
| Maxilla | 253 (55.24) | 22 (56.41) | 77 (52.74) | 81 (55.86) | 73 (57.03) |
| Mandible | 205 (44.76) | 17 (43.59) | 69 (47.26) | 64 (44.14) | 55 (42.97) |
| **Position, N (%)** |  |  |  |  |  |
| Anterior (canine-canine) | 83 (18.12) | 10 (25.64) | 19 (13.01) | 25 (17.24) | 29 (22.66) |
| Posterior | 375 (81.88) | 29 (74.36) | 127 (86.99) | 120 (82.76) | 99 (77.34) |
| **Side, N (%)** |  |  |  |  |  |
| Right | 234 (51.09) | 29 (74.36) | 74 (50.68) | 68 (46.90) | 63 (49.22) |
| Left | 224 (48.91) | 10 (25.64) | 72 (49.32) | 77 (53.10) | 65 (50.78) |
| **Type of Restoration, N (%)** |  |  |  |  |  |
| Single crown | 136 (29.69) | 8 (20.51) | 58 (39.73) | 49 (33.79) | 21 (16.41) |
| Bridge | 267 (58.30) | 17 (43.59) | 83 (56.85) | 82 (56.55) | 85 (66.41) |
| Overdenture | 14 (3.06) | 4 (10.26) | 0 (0.00) | 3 (2.07) | 7 (5.47) |
| Full-arch fixed restoration | 41 (8.95) | 10 (25.64) | 5 (3.42) | 11 (7.59) | 15 (11.72) |
| **Reason of Tooth Loss, N (%)** |  |  |  |  |  |
| Caries | 185 (40.39) | 14 (35.90) | 64 (43.84) | 56 (38.62) | 51 (39.84) |
| Periodontitis | 151 (32.97) | 14 (35.90) | 37 (25.34) | 53 (36.55) | 47 (36.72) |
| Trauma | 15 (3.28) | 0 (0.00) | 6 (4.11) | 1 (0.69) | 8 (6.25) |
| Agenesis | 8 (1.75) | 0 (0.00) | 5 (3.42) | 0 (0.00) | 3 (2.34) |
| Other reason/Unknown | 99 (21.62) | 11 (28.21) | 34 (23.29) | 35 (24.14) | 19 (14.84) |
| **Implant Length** (mm), mean (SD) | 9.91 (1.72) | 10.21 (1.90) | 9.73 (1.49) | 9.94 (1.87) | 10.01 (1.75) |
| **Implant Diameter** (mm), mean (SD) | 4.14 (0.41) | 4.06 (0.38) | 4.11 (0.40) | 4.22 (0.44) | 4.11 (0.39) |

Total number varies according to missing data for each variable.
Implant brands: S, Straumann; N, Nobel Biocare; A, AstraTech.
SD, standard deviation; N, number.

### Patient perception about peri-implant health status

Table 3 depict the patient perception about the health status of their implants. Although the implant-level prevalence of peri-implant health was only 8.52%, 91.67% of the implants were perceived as in health. A non-statistically significant tendency towards an increase in patient perception of disease with increasing disease severity was observed. However, even in case of peri-implantitis, 88.89% of the implants were perceived as in health.

**Table 3.** Patient perception about peri-implant health and diseases.

| Patient perception about each implant | Overall (N=456*) | Peri-implant health (N=39) | Peri-implant mucositis (N=146) | Pre-periimplantitis (N=145) | Peri-implantitis (N=126) | Healthy vs. diseased** | Pre-periimplantitis/Peri-implantitis vs. healthy or mucositis | Peri-implantitis vs. no peri-implantitis |
| --- | --- | --- | --- | --- | --- | --- | --- | --- |
| **'Do you think that your implant is in health?', N (%)** |  |  |  |  |  |  |  |  |
| No | 38 (8.33) | 1 (2.56) | 10 (6.85) | 13 (8.94) | 14 (11.11) | p=0.380 | p=0.421 | p=0.956 |
| Yes | 418 (91.67) | 38 (97.44) | 136 (93.15) | 132 (91.03) | 112 (88.89) |  |  |  |

N, number of implants.
\* missing data for 2 implants.
\*\* diseased implants included peri-implant mucositis, pre-periimplantitis and peri-implantitis.

### OHIP-14Sp according to peri-implant health and diseases

The oral health impact profile-14 according to peri-implant health and diseases is reported in table 4. The lower mean value was observed for handicap (0.33), while the highest for physical pain (2.62). The OHIP-14Sp mean sum score was 7.84, indicating a low level of oral health related impairment. No differences were observed among the different peri-implant health status, with the exception of physical pain, which was lower in patients affected by peri-implantitis (p=0.014).

**Table 4.** Oral health impact profile-14 according to peri-implant health and diseases.

| Oral Health Impact Profile-14 | Overall<br>(N=99) | Peri-implant<br>health/mu-<br>cositis<br>(N=12) | Pre-<br>periimplan-<br>titis<br>(N=31) | Peri-implantitis<br>(N=56) | *p-value* |
| --- | --- | --- | --- | --- | --- |
| **OHIP-14**, mean (SD) |  |  |  |  |  |
| Functional limitation | 0.91 (1.07) | 1.08 (0.99) | 0.97 (1.01) | 0.86 (1.13) | p=0.417 |
| Physical pain | 2.62 (1.69) | 3.08 (1.31) | 3.12 (1.62) | 2.25 (1.72) | p=0.014* |
| Psychological discomfort | 1.93 (2.10) | 1.83 (2.03) | 1.87 (2.16) | 2 (2.11) | p=0.975 |
| Physical disability | 0.45 (1.00) | 0.42 (1.16) | 0.39 (0.95) | 0.5 (1.01) | p=0.847 |
| Psychological disability | 1.16 (1.53) | 1.41 (1.44) | 1.12 (1.35) | 1.12 (1.66) | p=0.475 |
| Social disability | 0.41 (1.01) | 0.5 (0.90) | 0.45 (1.29) | 0.37 (0.86) | p=0.859 |
| Handicap | 0.33 (0.89) | 0.33 (0.89) | 0.19 (0.65) | 0.41 (1.00) | p=0.133 |
| **OHIP-14 Total**, mean (SD) | 7.84 (6.30) | 8.66 (4.41) | 8.12 (6.43) | 7.51 (6.63) | p=0.534 |

N, number of implants; SD, standard deviation.
\* p<0.05.

### Symptoms of peri-implant diseases

The frequency distribution of potential symptoms according to peri-implant health status is reported in table 5. History of pain was experienced, at least occasionally, on 7.64% of the implants, while history of spontaneous discomfort on 13.10%, history of bleeding on 14.63%, history of suppuration on 1.97%, history of swelling on 5.02% and discomfort during brushing on 15.72%. While the differences were no statistically significant, a clear tendency towards increased history of spontaneous discomfort, bleeding, swelling and discomfort during brushing was observed between implants with peri-implant diseases (PID) (peri-implant mucositis, pre-periimplantitis and peri-implantitis) and healthy peri-implant tissues. Indeed, spontaneous discomfort was present in around 14% of implants with PID, while in 2.59% of the healthy ones, history of bleeding in 10-22% of implants with PID vs. 5.13% of the healthy ones, history of swelling in around 5% of PID vs. 2.56% of the healthy ones; discomfort during brushing in around 16% of PID vs. 7.69% of the healthy ones. Only slight differences in frequency distributions among the different diseases (peri-implant mucositis vs. pre-periimplantitis vs. peri-implantitis) were observed for most of the symptoms. Even in presence of higher frequency of symptoms in disease vs. health, only a restricted number of implants with PID presented symptoms.

**Table 5.** Symptoms in peri-implant health and diseases.

| Symptoms about each implant | Overall<br>(N=458) | Peri-implant<br>health<br>(N=39) | Peri-implant<br>mucositis<br>(N=146) | Pre-<br>periimplan<br>titis<br>(N=145) | Peri-implantitis<br>(N=128) | Healthy vs.<br>diseased* | Pre-<br>periimplan<br>titis/Peri-implantitis<br>vs. healthy<br>or<br>mucositis | Peri-implantitis<br>vs. no peri-implantitis |
| --- | --- | --- | --- | --- | --- | --- | --- | --- |
| History of Pain, N (%) |  |  |  |  |  |  |  |  |
| Never | 423 (92.36) | 37 (94.87) | 132 (98.41) | 134 (92.41) | 120 (93.75) | p=0.673 | p=0.523 | p=0.412 |
| At least occasionally | 35 (7.64) | 2 (5.13) | 14 (9.59) | 11 (7.59) | 8 (6.25) |  |  |  |
| History of Spontaneous Discomfort, N (%) |  |  |  |  |  |  |  |  |
| Never | 398 (86.90) | 38 (97.44) | 127 (86.99) | 123 (84.83) | 110 (85.94) | p=0.121 | p=0.329 | p=0.922 |
| At least occasionally | 60 (13.10) | 1 (2.59) | 19 (13.01) | 22 (15.17) | 18 (14.06) |  |  |  |
| History of Bleeding, N (%) |  |  |  |  |  |  |  |  |
| Never | 391 (85.37) | 37 (94.87) | 113 (77.40) | 126 (86.90) | 115 (89.84) | p=0.215 | p=0.089 | p=0.237 |
| At least occasionally | 67 (14.63) | 2 (5.13) | 33 (22.60) | 19 (13.10) | 13 (10.16) |  |  |  |
| History of Suppuration, N (%) |  |  |  |  |  |  |  |  |
| Never | 449 (98.03) | 38 (97.44) | 142 (97.26) | 141 (97.24) | 128 (100.00) | p=0.526 | p=0.544 | NE |
| At least occasionally | 9 (1.97) | 1 (2.56) | 4 (2.74) | 4 (2.76) | 0 (0.00) |  |  |  |
| History of Swelling, N (%) |  |  |  |  |  |  |  |  |
| Never | 435 (94.98) | 38 (97.44) | 136 (93.15) | 140 (96.55) | 121 (94.53) | p=0.627 | p=0.476 | p=0.869 |
| At least occasionally | 23 (5.02) | 1 (2.56) | 10 (6.85) | 5 (3.45) | 7 (5.47) |  |  |  |
| Discomfort During Brushing, N (%) |  |  |  |  |  |  |  |  |
| Never | 386 (84.28) | 36 (92.31) | 129 (82.19) | 122 (84.71) | 108 (84.38) | p=0.419 | p=0.830 | p=0.922 |
| At least occasionally | 72 (15.72) | 3 (7.69) | 26 (17.81) | 23 (15.86) | 20 (15.62) |  |  |  |

N, number of implants; NE, not estimable.
\* diseased implants included peri-implant mucositis, pre-periimplantitis and peri-implantitis.

### Clinical signs of peri-implant diseases

The distribution of potential signs of peri-implant diseases is reported in table 6, while a representative study clinical case is reported in Figure 1.

**Table 6.** Clinical signs of peri-implant health and diseases.

| Signs regarding each implant | Overall (N=458) | Peri-implant health (N=39) | Peri-implant mucositis (N=146) | Pre-periimplantitis (N=145) | Peri-implantitis (N=128) | Healthy vs. diseased*<br>*** | Pre-periimplantitis/<br>Peri-implantitis vs.<br>healthy or mucositis | Peri-implantitis vs.<br>no peri-implantitis |
| --- | --- | --- | --- | --- | --- | --- | --- | --- |
| **Redness, N (%)** |  |  |  |  |  |  |  |  |
| Not at all | 101 (22.05) | 8 (20.51) | 38 (26.03) | 33 (22.76) | 22 (17.19) | *ref* | *ref* | *ref* |
| Mild | 241 (52.62) | 26 (66.67) | 76 (52.05) | 70 (48.28) | 69 (53.91) | p=0.367 | p=0.732 | p=0.418 |
| Moderate | 99 (21.62) | 5 (12.82) | 28 (19.18) | 36 (24.83) | 30 (23.44) | p=0.807 | p=0.149 | p=0.421 |
| Severe | 17 (3.71) | 0 (0.00) | 4 (2.74) | 6 (4.14) | 7 (5.47) | NE | p=0.584 | p=0.622 |
| **Swelling, N (%)** |  |  |  |  |  |  |  |  |
| Not at all | 190 (41.48) | 20 (51.28) | 62 (42.47) | 54 (37.24) | 54 (42.19) | *ref* | *ref* | *ref* |
| Mild | 220 (48.03) | 18 (46.15) | 70 (47.95) | 71 (48.97) | 61 (47.66) | p=0.991 | p=0.971 | p=0.419 |
| Moderate | 44 (9.61) | 1 (2.56) | 14 (9.59) | 17 (11.72) | 12 (9.38) | p=0.154 | p=0.242 | p=0.786 |
| Severe | 4 (0.87) | 0 (0.00) | 0 (0.00) | 3 (2.07) | 1 (0.78) | NE | NE | p=0.581 |
| **Deepest PPD, mean (SD)** | 5.05 (1.53) | 4.00 (0.88) | 5.13 (1.32) | 5.13 (1.42) | 5.21 (1.90) | p=0.000<br>*** | p=0.050 | p=0.154 |
| **Deepest PPD, N (%)** |  |  |  |  |  |  |  |  |
| ≤3 mm | 58 (12.99) | 9 (23.08) | 11 (7.53) | 17 (11.72) | 21 (16.41) | p=0.509 | p=0.215 | p=0.314 |
| 4-5 mm | 245 (53.49) | 28 (71.79) | 84 (57.53) | 76 (52.41) | 57 (44.53) | *ref* | *ref* | *ref* |
| 6-7 mm | 124 (27.07) | 2 (5.13) | 45 (30.82) | 41 (28.28) | 36 (28.12) | p=0.009<br>** | p=0.1113 | p=0.142 |
| ≥8 mm | 31 (6.77) | 0 (0.00) | 6 (4.11) | 11 (7.59) | 14 (10.94) | NE | p=0.008** | p=0.057 |
| **Deepest Mucosal Dehiscence, mean (SD)** | 0.38 (0.78) | 0.5 (0.82) | 0.20 (0.51) | 0.32 (0.65) | 0.62 (1.04) | p=0.590 | p=0.042* | p=0.005** |
| **Mucosal Dehiscence (at least 1 site), N (%)** |  |  |  |  |  |  |  |  |
| 0 mm | 345 (75.33) | 26 (66.67) | 122 (83.56) | 111 (76.55) | 86 (67.19) | *ref* | *ref* | *ref* |
| 0.5-1 mm | 62 (13.54) | 7 (17.95) | 18 (12.33) | 21 (14.48) | 16 (12.50) | p=0.278 | p=0.817 | p=0.517 |
| >1 mm | 51 (11.14) | 6 (15.38) | 6 (4.11) | 13 (8.97) | 26 (20.31) | p=0.529 | p=0.036* | p=0.006** |
| **Deepest PPD+Dehiscence, mean (SD)** | 5.18 (1.54) | 4.26 (1.02) | 5.18 (1.36) | 5.18 (1.38) | 5.46 (1.91) | p=0.001<br>*** | p=0.033* | p=0.026* |
| **Deepest PPD+Dehiscence, N (%)** |  |  |  |  |  |  |  |  |
| ≤3 mm | 48 (10.48) | 8 (20.51) | 11 (7.53) | 14 (9.66) | 15 (11.72) | *ref* | *ref* | *ref* |
| 4-5 mm | 247 (53.93) | 27 (69.23) | 83 (56.85) | 79 (54.48) | 58 (45.31) | p=0.468 | p=0.554 | p=0.714 |
| 6-7 mm | 129 (28.17) | 4 (10.26) | 45 (30.82) | 41 (28.28) | 39 (30.47) | p=0.020* | p=0.726 | p=0.425 |
| ≥8 mm | 34 (7.42) | 0 (0.00) | 7 (4.79) | 11 (7.59) | 16 (12.50) | NE | p=0.079 | p=0.125 |
| **Extent of BoP** |  |  |  |  |  |  |  |  |
| 0 sites/implant | 39 (8.52) | 39 (100.00) | 0 (0.00) | 0 (0.00) | 0 (0.00) | NE | NE | NE |
| 1-2 sites/implant | 129 (28.17) | 0 (0.00) | 42 (28.77) | 44 (30.34) | 43 (33.59) | *ref* | *ref* | *ref* |
| 3-4 sites/implant | 166 (36.24) | 0 (0.00) | 52 (35.62) | 67 (46.21) | 47 (36.72) | NE | p=0.394 | p=0.635 |
| 5-6 sites/implant | 124 (27.07) | 0 (0.00) | 52 (35.62) | 34 (23.45) | 38 (29.69) | NE | p=0.177 | p=0.847 |
| **Presence of SoP** |  |  |  |  |  |  |  |  |
| 0 sites/implant | 415 (90.61) | 39 (100.00) | 128 (87.67) | 133 (91.72) | 115 (89.84) | *ref* | *ref* | *ref* |
| At least 1<br>site/implant | 43 (9.39) | 0 (0.00) | 18 (12.33) | 12 (8.28) | 13 (10.16) | NE | p=0.738 | p=0.719 |

N, number of implants; SD, standard deviation; NE, not estimable.
\* p<0.05.
\*\* p<0.01.
\*\*\* p<0.001.
\*\*\*\* diseased implants included peri-implant mucositis, pre-periimplantitis and peri-implantitis.

**Figure 1.**
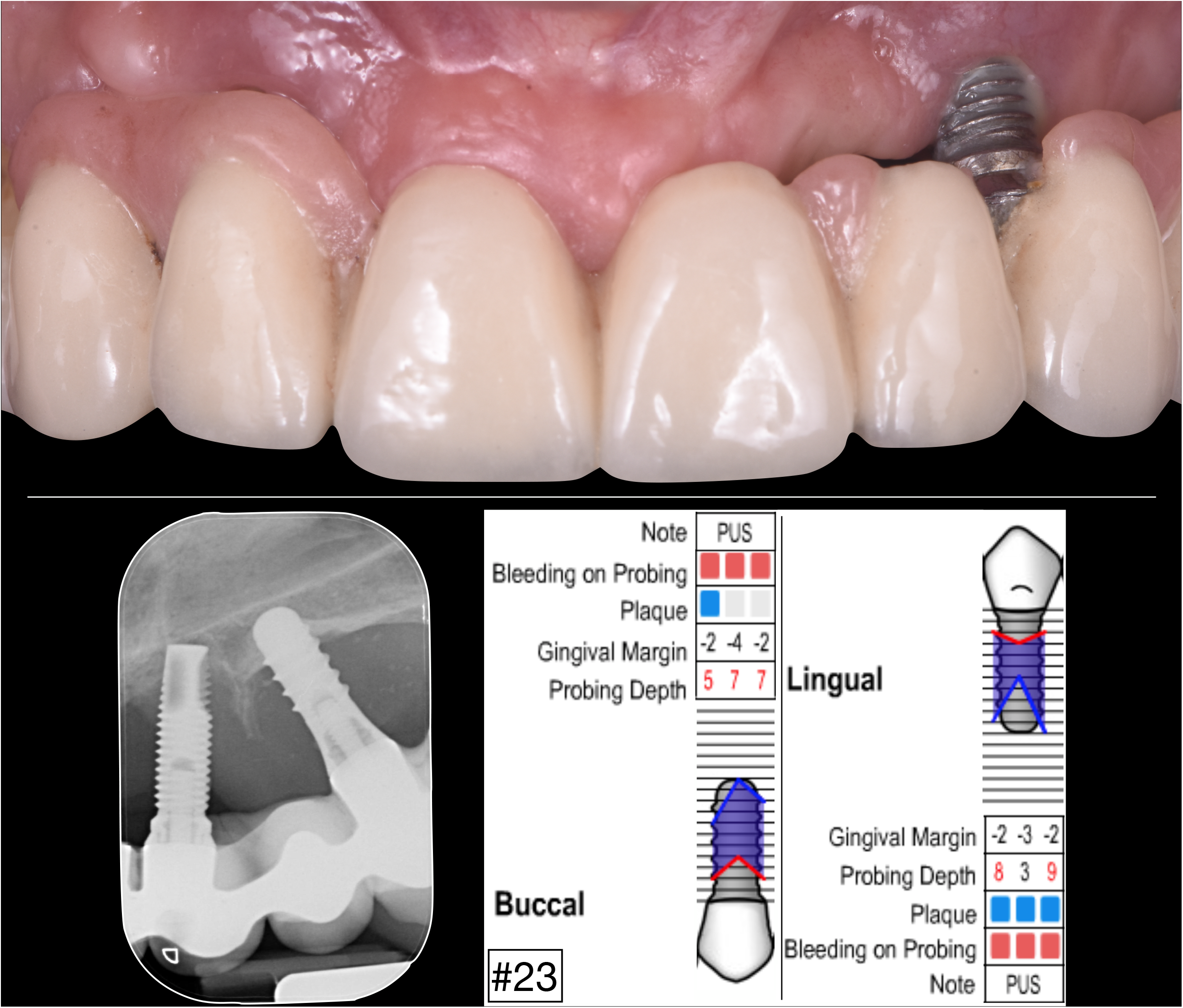
Representative study clinical case about clinical signs of peri-implantitis. Footnote: A study case of peri-implantitis presenting without visible swelling and redness, but with deep PPD, mucosal dehiscence and SoP (periodontal chart reported using: http://www.periodontalchart-online.com/).

Severe redness was only observed in presence of PID; also increased frequencies of moderate/severe swelling were more frequently detected in PID (around 10%) than in health (2.56%). However, for these signs, differences among the PID were not statistically significant. Similarly, while by definition the presence of BoP and SoP was only present in implants with PID, the extent of BoP and the presence of SoP did not differ between the different diseases.

Mean PPD was higher in implants with PID than in the healthy ones (p<0.001), while no differences were observed between different diseases. PPD≥6 mm was more frequent in PID (35%-40%) than in healthy implants (5.13%) (p<0.01), while PPD≥8 in pre-periimplantitis/peri-implantitis (8-11%) than in health/mucositis (0%-4%) (p<0.01).

The presence of mucosal dehiscence exceeding 1 mm was higher in health (15.38%) and in peri-implantitis (20.31%) than in peri-implant mucositis (4.11%) and pre-periimplantitis (8.97%). In particular, implants with peri-implantitis had a statistically significant higher mean mucosal dehiscence than implants without peri-implantitis (p<0.01).

## Discussion

This cross-sectional investigation on a representative sample from patients treated in a university postgraduate clinic has shown that peri-implant pathology is in most cases asymptomatic and leads to silent diseases. Even if a slight tendency to higher perception of disease was observed with increased PID severity, most of the implants were perceived as healthy, regardless of their true health status. Indeed, almost 90% of the peri-implantitis implants were considered as healthy by the patients.

Similarly, the impact of peri-implant diseases on patients’ oral health quality of life was very small, contrarily to what happen for periodontitis (Durham et al., 2013). This difference may be explained by the presence, in periodontitis cases, of a more mouth-generalized pathology, which in the most advanced cases also involves tooth mobility and masticatory disfunction, which in peri-implantitis only happen after the loss of osteointegration (implant loss). Peri-implantitis patients, in spite of having a negligible impact in their perceived oral health related quality of life, reported lower levels for the domain “physical pain”. This fact may be due to the characteristics of this university sample, where peri-implantitis was mostly present in patients also suffering moderate/severe periodontitis (Romandini et al., 2020), thus dentinal hypersensitivity (which define one of the two OHIP-14Sp questions regarding “physical pain”) may be reduced in those patients due to the reduced number of remaining teeth.

In the same direction of scarce perception of the disease by the affected patients, only a minor part of diseased implants presented with symptoms. While in periodontitis some symptoms (i.e. tooth mobility or bleeding during toothbrushing - Chatzopoulos et al., 2018; Dietrich et al., 2007; Eke et al., 2013) could alert patients to seek dental care before the disease is too advanced, for peri-implant diseases this appeared not to be the case. This highlights once more the importance of regular maintenance recalls, not only as a preventive measure, but as unmissable occasion for regular screening of patients with dental implants to allow for early diagnosis/intervention.

During these maintenance recalls, it is important to have sensitive clinical signs to recognize the diseases in their early stages. The presence of bleeding on probing is an important tool, but - beside its high sensitivity in identifying peri-implant diseases (since disease is defined by its presence) - it has a very low specificity in identifying which implants have peri-implantitis. Mucosal redness or swelling, extent of BoP and presence of suppuration, were not associated to specific peri-implant diagnosis. Only PPD, presence of mucosal dehiscence and their combinations were the clinical signs associated with pre-periimplantitis/peri-implantitis. However, while increased PPD was only associated with PID, the presence of mucosal dehiscence was associated with both peri-implantitis and peri-implant health. Consequently, mucosal dehiscence appeared to be the clinical sign which better identify the presence of peri-implantitis only when associated with the presence of BoP/SoP. These findings suggest that bone loss due to peri-implantitis may happen associated either to an increase in PPD or to a mucosal dehiscence, thus limiting the utility of increases in PPD as the sole clinical parameter to suspect the presence of bone loss in all cases. An increase in mean mucosal recession during peri-implantitis is also supported by an experimental study in beagle dogs (Monje, Insua, et al., 2018b). Indeed, the combination of PPD with mucosal dehiscence increased in the present study the diagnostic value of the single clinical signs alone.

When comparing the present results with the available literature, there is seldom information on the patient perception about peri-implant diseases and their impact on oral health quality of life. A qualitative study on patients with peri-implantitis reported how several patients had no perception of being diseased (Abrahamsson, Wennström, Berglundh, & Abrahamsson, 2017). Insua et al. (Insua, Monje, Wang, & Inglehart, 2017) reported an impaired OHRQoL in patients with peri-implantitis, however, in this cohort the patients had prior awareness of their peri-implant diagnosis. Increased PPD has been previously related to the presence of bone loss (Serino, Turri, & Lang, 2013) and of peri-implantitis (Monje, Caballé-Serrano, et al., 2018a; Monje, Insua, et al., 2018b; Ramanauskaite et al., 2018; Rodrigo et al., 2018; Vignoletti et al., 2019). However, the diagnostic value of PPD has been questioned, since cases of peri-implantitis may manifest with shallow PPD (Fransson, Wennström, & Berglundh, 2008) and accurate peri-implant probing is difficult to perform in presence of implant-supported restorations (Serino et al., 2013).

Contrarily to the present findings, Ramanauskaite et al. (Ramanauskaite et al., 2018) reported a relationship between the extent of BoP (number of bleeding sites/implant) and the presence of peri-implantitis, while Monje et al. (Monje, Caballé-Serrano, et al., 2018a) associated the presence of redness with peri-implantitis. SoP has been reported in different cohorts to be present only in a minority (10%-20%) of implants with peri-implantitis (Monje, Caballé-Serrano, et al., 2018a; Ramanauskaite et al., 2018; Rodrigo et al., 2018), which is in agreement with the findings of the present study. However, in the present study, similarly to Monje et al. (Monje, Caballé-Serrano, et al., 2018a) and Rodrigo et al. (Rodrigo et al., 2018), SoP was not a pathognomonic finding of peri-implantitis as it was present also in some peri-implant mucositis cases. Contrarily, Ramanauskaite et al. (Ramanauskaite et al., 2018) only found SoP on implants affected by peri-implantitis.

The present cross-sectional study, analysing a university representative sample with 458 implants, has evaluated the patient-reported perception and symptoms associated with presence of peri-implant diseases, as well as their impact on oral health related quality of life and the presence of clinical signs affecting the available dental implants. Both patients and examiners were blinded regarding the peri-implant health status, favouring the true patient’s perception, symptoms, OHRQoL and the results of the clinical evaluation. However, as most of studies reported in the literature, the absence of baseline documentation did not allow to identify peri-implantitis through direct evidence, what may lead to misclassification bias. Another limitation was the impossibility to obtain trusty information regarding previous peri-implantitis treatment (Ravidà, Galli, et al., 2020a), which may have an impact of patient perception, symptoms and signs of disease. Finally, even if representative from a university clinic, the present results suffer from limited generalizability, since most of these patients also suffered from periodontal diseases.

## Conclusions

Peri-implant diseases are in most cases asymptomatic and not perceived by the patients, what may prevent them to seek for dental care before the disease is too advanced. Characteristic signs of peri-implant disease, such as SoP, the extent of BoP or mucosal redness, were not associated to a specific peri-implant diagnosis, while increased PPD, presence of mucosal dehiscence and their combination were associated with pre-periimplantitis/peri-implantitis. However, there is no clinical parameter really able to discriminate between implants affected by peri-implant mucositis and peri-implantitis. Prospective cohort studies are needed to evaluate whether changes in clinical parameters, more than their one-time assessment, are able to identify peri-implantitis or at least to give specific indications for radiographic evaluation. Moreover, research is needed to study the potential role of new diagnostic technologies to improve the diagnosis of peri-implantitis in its early stages.

## Data Availability

Upon request, data can be shared.

## Acknowledgments

The authors wish to kindly thank Mariano del Canto, Laura Sobrino and Javier Calatrava for their kind support during part of the data collection process. Moreover, the authors are grateful to Cristiano Tomasi and Anna Chiara Frigo for their kind statistical advices.

The authors declare no conflicts of interest related to this study. This study was partially funded by the Osteology Foundation through a Young Research Grant to Dr. Mario Romandini (project n. 15-251).

## Author contributions

M.R. conceived the ideas; C.L., I.P., A.A., M.C.S. and M.R. collected the data; M.R. and M.C.S. and M.S. analysed the data; and M.R. and M.S. led the writing.

